# Comparison of severe and non-severe COVID-19 pneumonia: review and meta-analysis

**DOI:** 10.1101/2020.03.04.20030965

**Authors:** Weiping Ji, Jing Zhang, Gautam Bishnu, Xudong Du, Xinxin Chen, Hui Xu, Xiaoling Guo, Zhenzhai Cai, Xian Shen

## Abstract

**Objective:** To compare the difference between severe and non-severe COVID-19 pneumonia and figure out the potential symptoms lead to severity.

**Methods:** Articles from PubMed, Embase, Cochrane database, and google up-to 24 February 2020 were systematically reviewed. Eighteen Literatures were identified with cases of COVID-19 pneumonia. The extracted data includes clinical symptoms, age, gender, sample size and region et al were systematic reviewed and meta analyzed.

**Results:** 14 eligible studies including 1,424 patients were analyzed. Symptoms like fever (89.2%), cough (67.2%), fatigue (43.6%) were common, dizziness, hemoptysis, abdominal pain and conjunctival congestion/conjunctivitis were rare. Polypnea/dyspnea in severe patients were significantly higher than non-severe (42.7% vs.16.3%, P<0.0001). Fever and diarrhea were higher in severe patients(p=0.0374and0.0267). Further meta-analysis showed incidence of fever(OR1.70,95%CI 1.01-2.87), polypnea/dyspnea(OR3.53, 95%CI 1.95-6.38) and diarrhea(OR1.80,95%CI 1.06-3.03) was higher in severe patients, which meant the severe risk of patients with fever, polypnea/dyspnea, diarrhea were 1.70, 3.53, 1.80 times higher than those with no corresponding symptoms.

**Conclusions:** Fever, cough and fatigue are common symptoms in COVID-19 pneumonia. Compared with non-severe patients, the symptoms as fever, polypnea/dyspnea and diarrhea are potential symptoms lead to severity.

## Introduction

In December 2019, a series of unknown caused pneumonia appeared in Wuhan, it’s clinical manifestations were very similar to viral pneumonia^[1]^. Later novel coronavirus was found as source virus, and named COVID-19 ^[2]^. At present, novel coronavirus pneumonia (NCP) had been rapidly spread throughout China and overseas countries ^[3-5]^. Human-to-human transmission of this virus was proved ^[6-7]^. The clinical features of NCP patients were diversified, in severe cases it may lead to acute respiratory distress syndrome (ARDS) and even death^[8-11]^. Currently, no systematic review and Meta-analysis of the clinical characteristics of NCP was found. This paper reviewed and compared the difference between severe and non-severe of the disease, which would help to better understand and guide future.

## Method

### Sources and search criteria

We conducted a comprehensive systematic search of PubMed, Embase, Cochrane database, and google to find all published studies that describe the clinical characteristics of COVID-19, using the search terms, “novel coronavirus”, “SARS-CoV-2”, “COVID-19”. According to title and abstract, two independent researchers selected and classified literatures, and reviewed all followed criteria.

### Inclusion and exclusion criterina

Literatures described NCP patient’s clinical signs. If there were duplicates in the study population, the literature with the largest sample size will be included. Cases that did not describe the clinical characteristics of NCP patients and repeated cases were excluded. Articles published by the same author were carefully screened to identify duplication.

### Data extraction and analysis

All available publications have been carefully analyzed and strictly reviewed. The collected clinical data include population data (age, gender), number of patients and clinical manifestations. Full-text versions of relevant articles were reviewed, quality of the selected articles were evaluated, and clinical characteristics were extracted. We compared the clinical characteristics of severe and non-severe patients originally from three articles classified NCP patients as severe and non-severe.

### Statistical analysis

SAS 9.4 and Review Manager 5.3 software were used for analysis and drawing. Continuous variables were represented by median and quartile ranges (IQR). Classification variables were summarized as counts and percentages in each category. The age, gender, number and clinical signs of NCP patients were statistically described; Cochran-mantel-Haenszel test (stratified chi-square test) was used to compare the differences between severe and non-severe NCP. With OR (Odds ratio) as the effect quantity, we used Mantel-Haenszel test with fixed or random effect for further meta-analysis of the clinical signs with statistical differences, and showed by drawing forest map. Symmetry tested by funnel plot.

## Results

### Search result

A total of 333 relevant literatures published in PubMed and other databases were detected, 68 were removed because of duplication. 265 were removed based on the inclusion criteria. 23 literaturesdid not report clinical signs. 4 were excluded because of casesoverlapped[8-9.12-13]. Finally, 14 articles were included for the final analysis[10-11.14-25]. Fig 1 shows the study selection flowchart. Data from all eligible studies were obtained frompublished manuscripts.

**Figure 1:**
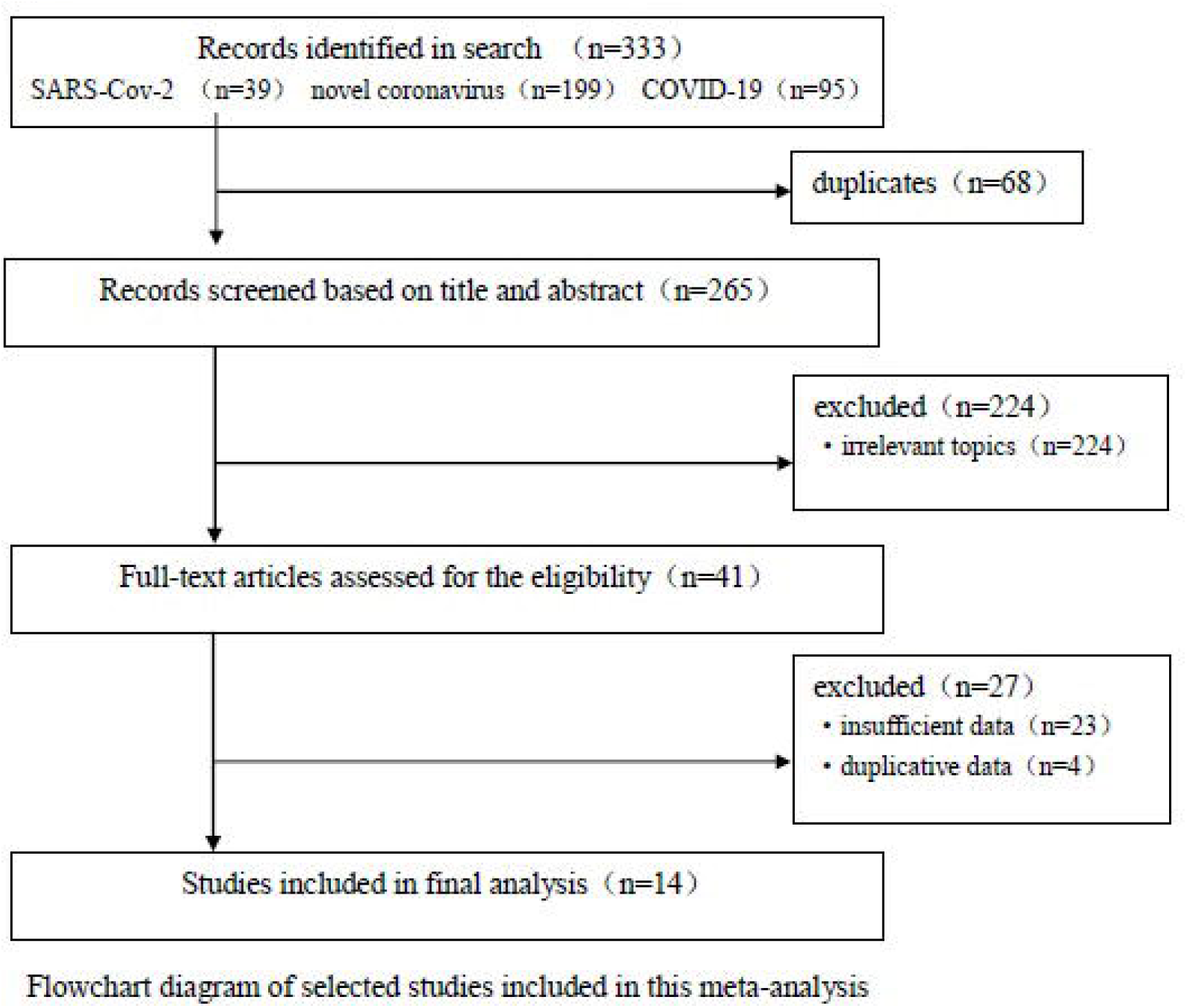
After selection, 5 papers of group research and 9 articles of case report were eligible, with a total of 1,424 cases. 3 papers with a total of 1,377 cases reported clinical signs of severe and non-severe patients subgroups (literature 2 was divided into ICU and non-ICU, which were assumed equivalent to severe and non-severe).

### Synthesis of results

A systematic review showed that 610 patients (42.8%) with NCP were female. Fever (89.2%) and cough (67.2%) were the most common symptoms, followed by fatigue (43.6%), phlegm (28.6%), shortness of breath/difficulty(21.7%), and the less common symptoms were dizziness, hemoptysis, abdominalpain, conjunctivalcongestion/conjunctivitis.1,377 cases were divided into severe group (1,110) and non-severe group (267), Stratified chi-square test showed that there was nosignificant difference in gender between the two groups(P>0.05), and the median age of severe patients was slightly older. Polypnea/dyspnea in severe patients were significantly higher than in non-severe patients (42.7% vs.16.3%, P<0.0001), fever and diarrhea were higher in severe patients (P=0.0374and0.0267). Conjunctival congestion/conjunctivitis (P=0.0176), hemoptysis (P=0.0344), anorexia (P=0.0008), dizziness (P=0.0023) and abdominal pain (P=0.0015) may highly occurred in severe patients, but the dates of these symptoms only been reported in one or two literatures, so inter-group comparisons of these symptoms should be treated with caution. (See fig 2 and 3)

**Figure 2:**
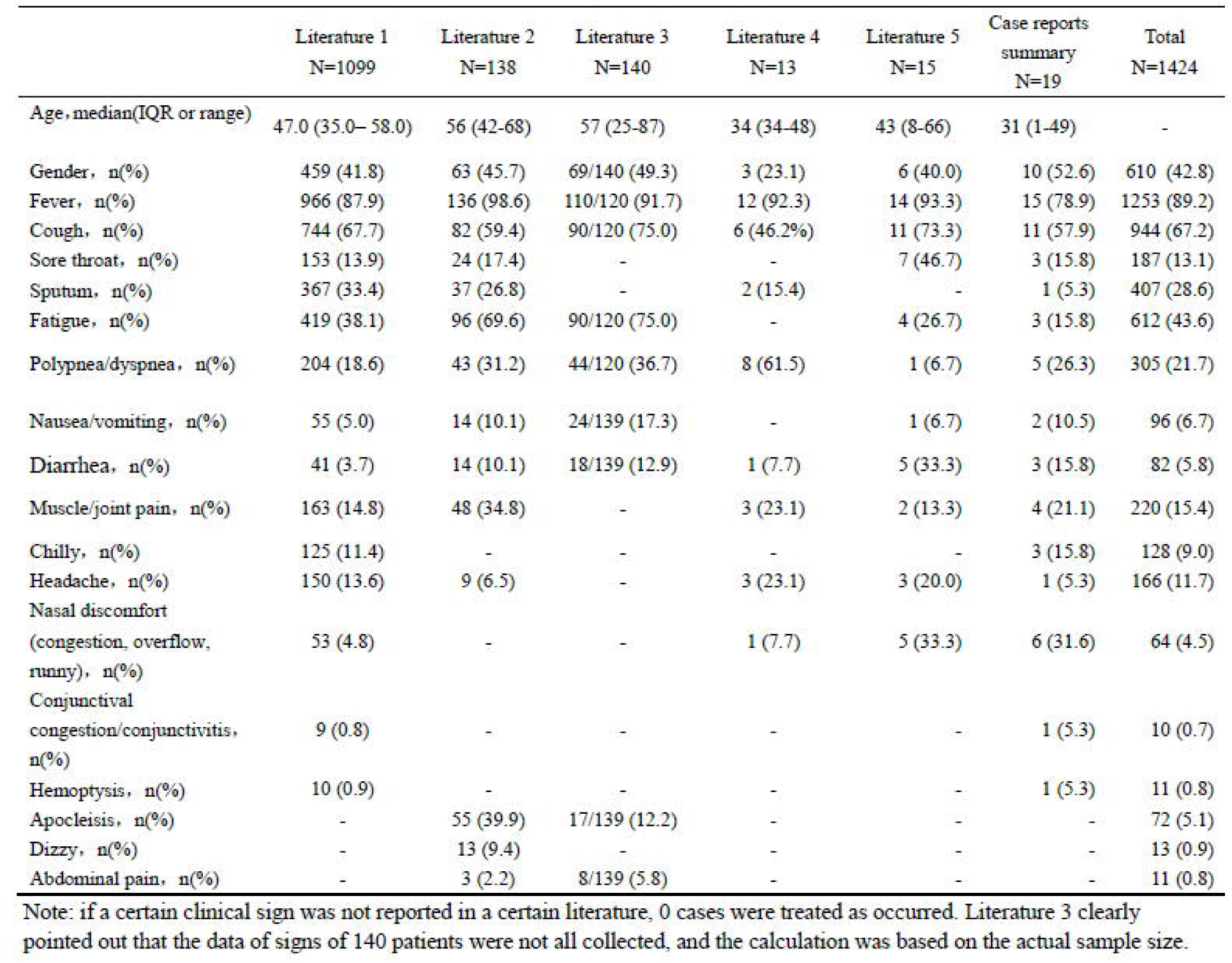
Fever, cough, polypnea/dyspnea and diarrhea were the most common clinical signs reported in the literature, while some other signs were more or less unreported. It assumed that if a certain clinical sign was not reported in a certain literature, it would be treated as none.

**Figure 3:**
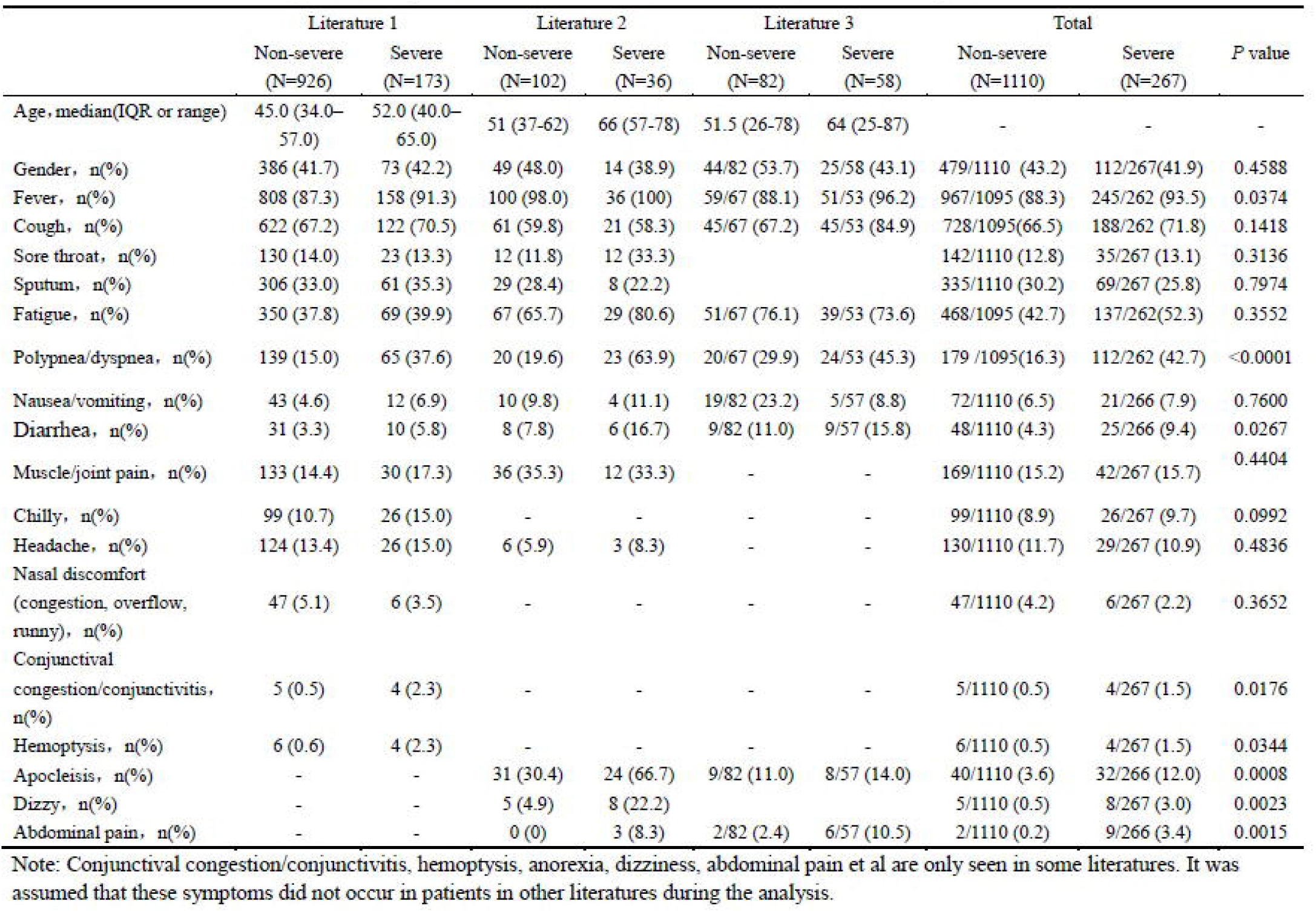
The comparison of clinical signs between severe and non-severe patients was shown among 1,377 cases, including 1,110 cases of non-severe patients and 267 cases of severe patients. Polypnea/dyspnea, fever and diarrhea showed significantly difference in two groups. In severe patients, conjunctival congestion/conjunctivitis, hemoptysis, anorexia, dizzinessand abdominal pain were more likely to occur, but because some data was incomplete, the results needed to be caution.

### Meta analysis results

3 literatures including 262 cases in the severe group (245 fever,93.51%) and 1,095 cases in the non-severe group (967 fever,88.31%) were used to meta analysis for fever; Three studies were considered to be homogeneous, and fixed effect model(Chi^2^=0.88, P=0.64, I^2^=0%). The pooled effect was 1.70 (95%CI, 1.01- 2.87) indicating fever patients had a 1.70 times higher incidence of severe risk than non-fever. On symptoms of polypnea/dyspnea, 262 cases in the severe group(112 polypnea/dyspnea,42.75%) and 1095 cases in the non-severe group(179polypnea/dyspnea,68.32%). Test showed that 3 literatures have certain heterogeneity(Chi2=5.26,P=0.07, I2=62%), so the random effect model was adopted; The individual OR effect showed differently, but combined OR effect was 3.53 (95%CI, 1.95-6.38), indicating the incidence of severe risk of the patients with polypnea/dyspnea was 3.53 times higher than those none. On symptoms of diarrhea, 266 cases in the severe group (25 diarrhea,9.40%) and 1,110 cases in the non-severe group (48diarrhea,4.32%). Test showed homogeneity (Chi^2^=0.32, P=0.85,I^2^= 0%) and fixed effect model was used, The combined OR value was 1.80 (95%CI, 1.06-3.03), indicating the incidence of diarrhea in the severe group significantly higher than non-severe group, and the severerisk of diarrheapatients was 1.80 times higher than those none. (See Fig 4)

**Figure 4:**
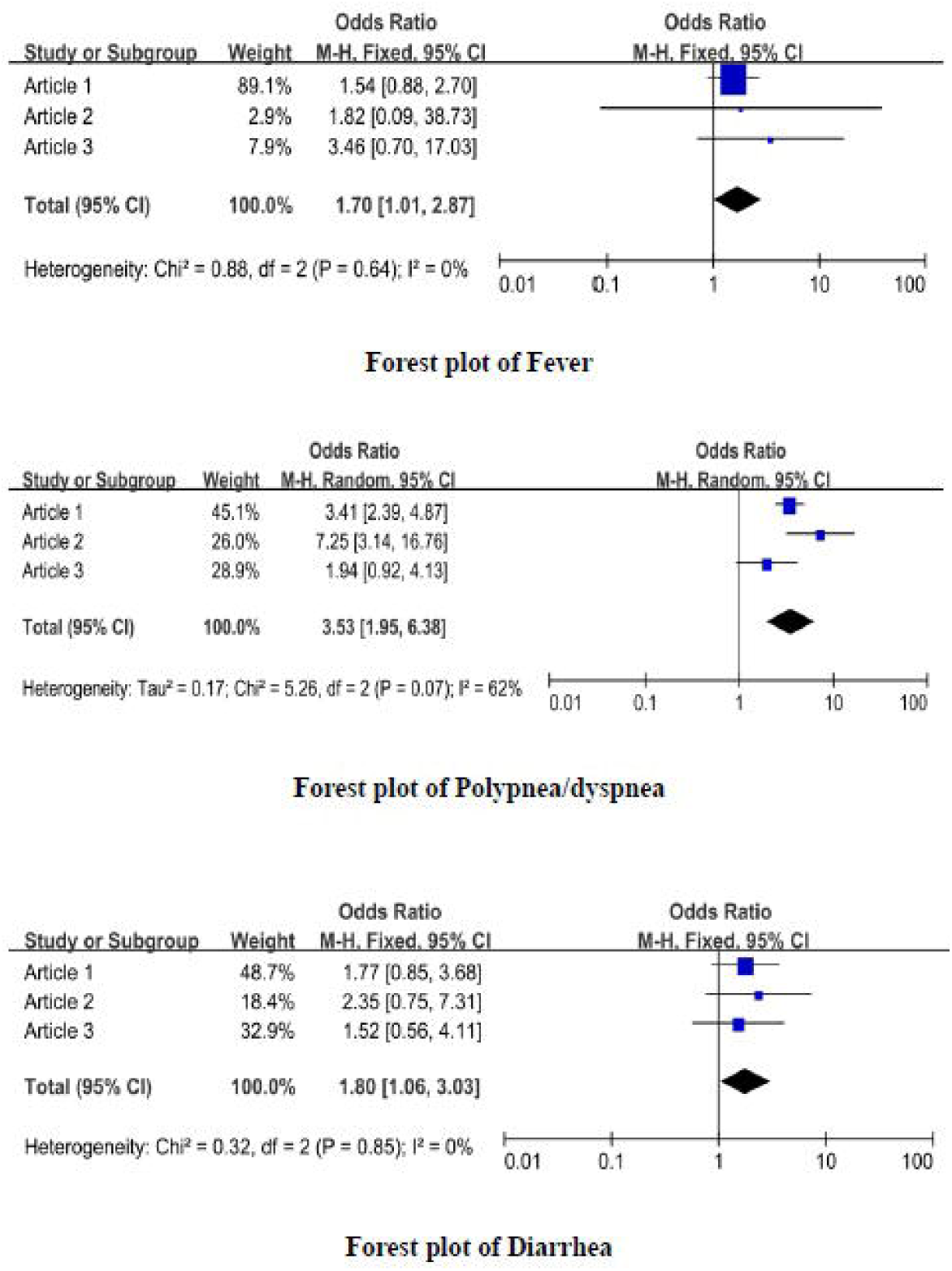
Further meta-analysis was performed for fever, polypnea/dyspnea, and diarrhea. Effect value OR analysis indicated severe risk of febrile, polypnea/dyspnea and diarrhea patients were 1.70 (95%CI, 1.01-2.87), 3.53 (95%CI, 1.95-6.38), 1.80 (95%CI, 1.06-3.03) times higher than those none.

The graphs in funnel plots of fever, polypnea/dyspnea and diarrhea are basically symmetrical, which looks like no significant publication deviation. (See fig 5)

**Figure 5:**
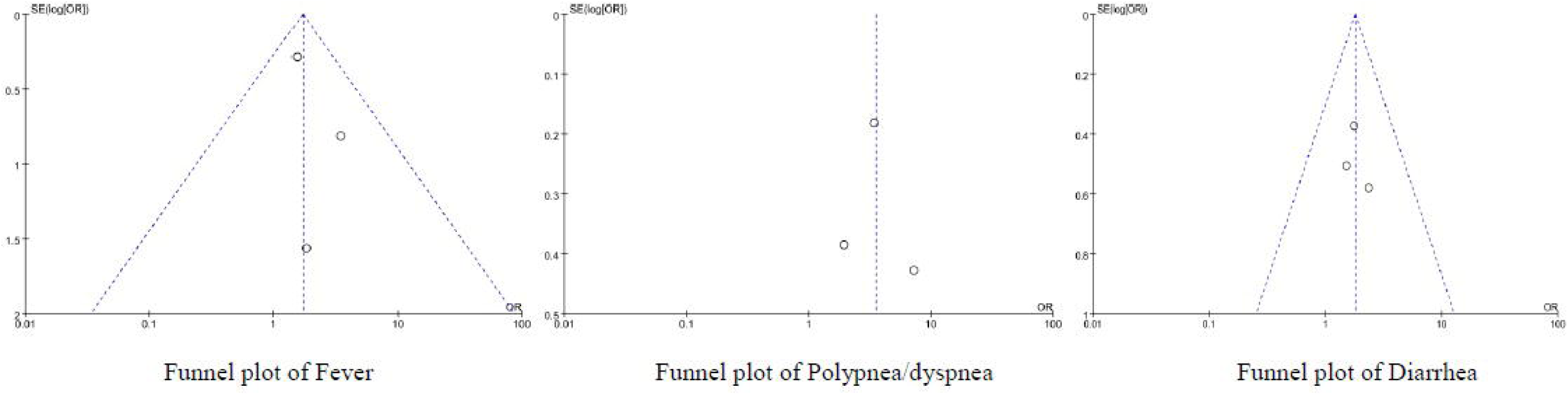
The graphs in funnel plots of fever, polypnea/dyspnea and diarrhea are basically symmetrical, which preliminary indicates no significant publication deviation.

## Discussion

There was no difference in proportion of gender between severe and non-severe NCP patients, which was consistent with the latest report[11-14]. Most of the patients showed fever and cough. Compared with non-severe patients, Fever, Polypnea/dyspnea and Diarrhea were more common in severe patients. But recent reports showed that there was no difference in the proportion of fever and diarrhea between severe and non-severe patients [10.11.14]. In addition, the symptoms of Polypnea/dyspnea were similar to those reports[10-11], but Zhang et al.[14]report had no statistical difference. One possible explanation was that in previous reports, the sample size of patients was relatively small or it came from the early epidemic phase of NCP.

NCP patients had the different clinical classification of disease severity. Non-severe NCP was with fever, respiratory tract symptoms, and image findings of pneumonia. Severe NCP met diagnostic criteria[26]. This review found preliminary evidence from three literatures that fever, polypnea/dyspnea and diarrhea are statistically significant diagnostic indicators for severe NCP patients and Polypnea/dyspnea was most predictable. Clinician had to arouse great attention on diagnostic value of them in future. In the early stages of NCP nonspecific signs and symptoms may be clinically indistinguishable from other common infectious diseases, especially winter respiratory virus. These characteristics of NCP had some similarities with SARS-CoVand MERS-CoVinfections[27.28]. However, patients with COVID-19 rarely had obvious signs and symptoms of upper respiratory tract (nasal obstruction, rhinorrhea, runny nose, sore throat). In addition, intestinal signs and symptoms such as diarrhea were rare in NCP patients, while about 20-25% of patients with MERS-CoV or SARS-CoV infection had diarrhea. It should be noted that fever of NCP(10.8%) patients was more common than SARS-CoV (1%) and MERS-CoV(2%)[29].

Our systematic review had limitations. Firstly, most of the data in this study are from retrospective studies and case reports, which usually report successful management and are affected by selection and publication bias. Secondly, the datacollection of some cases is incomplete. So the statistical test and the discovery of p-value should be carefully explained. Thirdly, the number of included studies is not enough, the test efficiency is insufficient, symmetry can be observed, but it is difficult to evaluate symmetry.

## Conclusion

The common symptoms NCP patients were fever, cough and fatigue. Compared with non-severe NCP patients, the symptoms as fever, polypnea/dyspnea and diarrhea were more common in severe patients. Those were potential symptoms which might lead patients to severity.

## Data Availability

All data referred to in the manuscript was obtained from online databases.

https://www.ncbi.nlm.nih.gov/pubmed

http://www.webofscience.com

http://www.clinicalkey.com

## Authors’ contributions

Weiping Ji, Jing Zhang, Hui Xu, Xiaoling Guo designed and completed the manuscript. Gautam Bishnu and Xudong Du completed data and language work, Zhenzhai Cai and Xinxin Chen completed statistics and verification, Xian Shen completed the review and revision of the paper, and guided the whole process.

## Acknowledgements

The authors declared no competing interests exist.

